# Age- and disease severity-associated changes in the nasopharyngeal microbiota of COVID-19 patients

**DOI:** 10.1101/2023.12.20.23300278

**Authors:** Fernando Pérez-Sanz, Sylwia D. Tyrkalska, Carmen Álvarez-Santacruz, Antonio Moreno-Docón, Victoriano Mulero, María L. Cayuela, Sergio Candel

## Abstract

Dysbiosis has been linked to the pathogenesis of multiple diseases. Although dozens of publications have associated changes in the nasopharyngeal microbiota to patient’s susceptibility to COVID-19, results from these studies are highly variable and contradictory in many cases. Addressing the limitations in previous research responsible for that variability, this study uses 16S rRNA gene sequencing to analyse the nasopharyngeal microbiota of 395 subjects, 117 uninfected controls and 278 COVID-19 patients, of different age groups that cover the entire lifespan and across varying disease severities. Importantly, our results reveal that bacterial diversity decreases progressively throughout life but only in severely ill COVID-19 patients, in whose nasopharynx, moreover, the opportunistic pathogen bacterial genera *Staphylococcus*, *Corynebacterium*, *Streptococcus*, *Prevotella*, *Acinetobacter*, and *Pseudomonas* are overrepresented. Notably, *Scardovia wiggsiae* appears only in severe COVID-19 patients over 60 years of age, suggesting a potential utility of this bacterial species as a COVID-19 severity biomarker in the elderly, who are the most susceptible individuals to suffer from serious forms of the disease and the age group that presents more differences in comparison with the other age groups according to the majority of the parameters analysed in this study. Our results provide valuable insights into age-associated dynamics within nasopharyngeal microbiota during severe COVID-19, offering potential avenues for further exploration and therapeutic interventions.

## BACKGROUND

The coronavirus disease 2019 (COVID-19) is caused by the novel betacoronavirus severe acute respiratory syndrome coronavirus 2 (SARS-CoV-2) (1), that penetrates the host through the upper airways (2). The COVID-19 outbreak, declared as a global pandemic by the World Health Organization on March 11, 2020 (3), has afflicted humans since its inception and continues to take a huge toll on human life and health, with almost 7 million deaths to date (https://covid19.who.int/ Accessed on November 16, 2023). Despite the striking efficacy that COVID-19 vaccines have shown so far (4, 5), the limited effectiveness of other treatments and the possibility of new variants emerging that circumvent the protection of such vaccines, requires a deeper knowledge of COVID-19 pathogenesis as well as the factors that make some human groups more susceptible to the disease, such as the elderly (6).

Among the different parts of the human upper respiratory tract, the nasopharynx is anatomically unique as it presents a common meeting place for the ear, nose, and mouth cavities (7). Because of this, nasopharyngeal epithelium plays a crucial role as a portal for initial infection and transmission of infectious droplet or aerosol-transmitted microorganisms, such as SARS-CoV-2 as demonstrated by the fact that nasopharyngeal swabs present higher viral loads than nasal (8), oropharyngeal (9), or throat (10) swabs. Thus, the study of the involvement of the nasopharynx in health and disease has gained a special prominence since the outbreak of the COVID-19 pandemic (11), and nasopharyngeal swabs are considered “gold standard” for the diagnosis of SARS-CoV-2 infections (8).

Microorganisms have been found to be part of the microbiota in the different locations of the healthy human body, where they form complex ecological communities and influence the human physiology (12). Even through the respiratory microbiota had not been so studied as that of other anatomical areas, such as the gut, due to the old paradigm that lungs were sterile (13, 14), recent studies have demonstrated that changes in the nasopharyngeal microbiota clearly correlate with increased or reduced susceptibility to different viral infections in humans (13). Indeed, focusing on COVID-19 research, dozens of studies have already tried to elucidate whether SARS-CoV-2 infection or the COVID-19 disease severity are associated with changes in the nasopharyngeal microbiota (14). However, unfortunately, the analysis of all these previous studies reveals extremely variable and contradictory results, which prevent solid and reliable conclusions from being drawn (14).

Given that the aforementioned variability could be mostly avoided as its possible sources have already been identified and discussed in depth (14), and the importance that the characterization of the correlations between changes in the nasopharyngeal microbiota and the infection by SARS-CoV-2 or the COVID-19 disease severity could have from a biomedical point of view, further research that provides new knowledge to this field while avoiding that variability is essential. Thus, the new pieces that are added to the still lacunar knowledge on nasopharyngeal microbiota and COVID-19 could open new therapeutic avenues to reduce the severity of COVID-19 patients and/or improve their disease outcome, for example with the strategy of manipulating the nasopharyngeal microbiota, which has already worked in the treatment of other diseases such as metabolic disorders, cancer, and other viral infections (15–17).

Here, we elude the majority of the previously identified potential sources of variability (14) and extend the study where we already characterized in detail the nasopharyngeal microbiota of healthy people throughout life (18), to now analyse the changes in such microbiota in COVID-19 patients with different disease severities and at all stages of life. For this, we analyse the diversity and relative abundance of the nasopharyngeal microbiota across the whole lifespan in a total of 395 individuals of all ages, both sexes, and with different COVID-19 disease severities, and the taxonomic changes in the nasopharynx associated to these parameters. We therefore provide a very comprehensive and valuable dataset that will allow to identify the possible relationships between changes in nasopharyngeal microbiota and susceptibility to or severity of COVID-19, with special interest in the most susceptible groups such as the elderly (6).

## METHODS

### Sample selection, collection, and classification

The uninfected nasopharyngeal control samples were selected, collected, and classified as previously described (18). Besides those uninfected samples, and according to our experimental design and economic resources, we decided to select and collect a maximum of 360 nasopharyngeal samples from SARS-CoV-2 infected individuals with different ages and COVID-19 disease severity, as will be detailed here after (Table S1). These samples were randomly selected from a cohort of 4,996 SARS-CoV-2 infected subjects belonging to the Health Area I of the Region of Murcia (Spain) who voluntarily provided their samples between 1 September 2020 and 3 November 2020 for diagnostic purposes and tested positive for SARS-CoV-2 infection. Nasopharyngeal swabs were obtained by approaching the nasopharynx transnasally and stored in Universal Transport Medium (UTM): Viral Transport medium (COPAN Diagnostics Inc., Murrieta, CA, USA). Nucleic acid extraction was performed using the automatized system Nuclisens EasymaG (bioMérieux, Madrid, Spain) based on the ability of silica to bind DNA and RNA in high salt concentrations (Boom technology). The polymerase chain reaction (PCR) kit used to verify that all the samples were positive for SARS-CoV-2 infection was Novel Coronavirus (2019-nCoV) Real Time Multiplex RT-PCR kit (Detection for 3 Genes), manufactured by Shanghai ZJ Bio-Tech Co., Ltd. (Liferiver Biotech, la Jolla, CA, USA) and the CFX96 Touch Real-Time PCR Detection System (BioRad, Madrid, Spain).

To facilitate the study of sex-, age, and COVID-19 disease severity-associated changes in the nasopharyngeal microbiota, and to ensure that the sample size of all the sex, age, and disease severity groups were homogeneous, we decided on an experimental design that distributed the maximum of 360 infected nasopharyngeal samples that we could analyse into three COVID-19 disease severity groups (mild, moderate, and severe) with a maximum of 120 individuals each, each divided into six age groups with 20 individuals each, of which 10 were females and the other 10 were males (Table S1). For this, the 4,996 SARS-CoV-2 infected individuals of our parent cohort were first divided into their COVID-19 disease severity matched groups, and later into their age matched groups within each of the severity groups and numbered. Then, randomly obtained numbers were used to select 10 females and 10 males from each of the age groups within each of the severity groups. Random numbers were generated in RANDOM.ORG, which is a True Random Number Generator (TRNG) that generates true randomness via atmospheric noise, unlike the most common and less trustworthy Pseudo-Random Number Generators (PRNGs) [RANDOM.ORG: True Random Number Service. Available at: https://www.random.org]. The reason that some groups contained fewer patients than planned in our experimental design, or even none such as patients under 20 years of age with moderate or severe COVID-19 disease severity, is that patients with these characteristics are rare and we simply enrolled all patients with such characteristics who were in our parent cohort (Table S1). Our COVID-19 disease severity groups were established according to the World Health Organization (WHO) COVID-19 severity classification [https://iris.who.int/bitstream/handle/10665/332196/WHO-2019-nCoV-clinical-2020.5-eng.pdf], with the only difference that the ‘severe disease’ and ‘critical disease’ groups established by the WHO are both gathered in our group of severe COVID-19. Finally, while the exclusion criteria for the uninfected nasopharyngeal samples were already described (18), the only SARS-CoV-2 infected individuals excluded from this study were those younger than 1 year as the microbiome of infants is known to be highly fluctuating with age and, therefore, it could significantly increase the variability of our analyses.

### Amplification, library preparation, and sequencing

Exactly as previously described for our uninfected control cohort (18).

### Bioinformatics and statistical analysis

The obtained sequences were analysed and annotated with the Ion Reporter 5.18.2.0 software (Thermo Fisher Scientific Inc., Alcobendas, Spain) using the 16S rRNA Profiling workflow 5.18. Clustering into OTUs and taxonomic assignment were performed based on the Basic Local Alignment Search Tool (BLAST) using two reference libraries, MicroSEQ® 16S Reference Library v2013.1 and the Greengenes v13.5 database. For an OTU to be accepted as valid, at least ten reads with an alignment coverage ≥ 90% between hit and query were required. Identifications were accepted at the genus and species level with sequence identity > 97% and > 99%, respectively. Annotated OTUs were then exported for analysis with R (v.4.1.2) (https://www.R-project.org/), where data were converted to phyloseq object (19) and abundance bar plots and heatmaps were generated. Data were converted to DESeq2 object (20), that uses a generalized linear model based on a negative binomial distribution, to calculate differential abundance between groups. Thus, the differential abundance analysis was conducted according to the phyloseq package vignette with bioconductor DESeq2 (https://bioconductor.org/packages/devel/bioc/vignettes/phyloseq/inst/doc/phyloseq-mixture-models.html#import-data-with-phyloseq-convert-to-deseq2, accessed on 16 November 2023). The raw abundance matrix was imported into phyloseq object (as specified in the documentation of phyloseq with DESeq2) and subsequently converted to DESeq2 object. Then, estimated size factors were used with the DESeq2 function to obtain the differential abundance. DESeq automatically searches for outliers and, if possible, replaces the outlier values estimating mean-dispersion relationship. If it’s not possible to replace, then p-values are replaced by NA. R (v.4.1.2) was also used to perform a principal coordinates analysis (PCoA) on Bray-Curtis dissimilarity measures among samples based on relative OTU abundances (i.e., percentages). The relative abundances of OTUs were also used to test for statistically significant differences between age groups. Group OTU compositions were compared through the non-parametric statistical tool ANOSIM. The 90% confidence data ellipses for each of the age groups were plotted. Alpha diversity was estimated based on Shannon and Inverse Simpson indices by using the phyloseq package. To test for statistically significant differences between age or COVID-19 disease severity groups in alpha diversity, the non-parametric Wilcoxon test was used. Heatmaps were generated by calculating the average abundance of each age group for each severity level. The values shown in the heatmaps were logarithmically transformed. The ‘heatmap.2’ function from the ‘gplots’ package in R was used for the visualization (https://CRAN.R-project.org/package=gplots, accessed on 16 November 2023). The bar plots aggregated by age groups show the aggregated relative abundance (sum of relative abundances). Krona charts, that aid in the estimation of relative abundances even within complex metagenomic classifications, were generated as previously described (21). All the other graphs were generated with the R package ggplot2 version 3.3.3., including the confidence data ellipses which were plotted using the ‘stat_ellipse’ function also from this package (22).

## RESULTS

### Data annotation

A total of 395 nasopharyngeal microbiomes from 117 uninfected control subjects and 278 SARS-CoV-2 infected individuals were analysed (Table S1). A total of 30,535,433 high quality 16S rRNA sequences ranging from 8,969 to 330,138 sequences per sample (mean = 77,305; median = 64,368) were obtained after quality control analyses and OTU filtering. The 16S rRNA sequences were binned into 167 families, 329 genera and 671 species. Considering all the samples together, the most abundant families were Moraxellaceae (26.4%), Pseudomonadaceae (15.8%), Prevotellaceae (12.0%), Streptococcaceae (11.0%) and Enterobacteriaceae (10.8%). The most abundant genera were *Acinetobacter* (28.2%), *Pseudomonas* (18.8%), *Streptococcus* (12.8%), *Prevotella* (11.9%) and *Brevundimonas* (6.5%). The most abundant species were *Acinetobacter johnsonii* (22.6%), *Prevotella melaninogenica* (19.0%), *Dolosigranulum pigrum* (13.4%), *Ralstonia pickettii* (9.0%) and *Brevundimonas halotolerans* (8.7%). To reveal any potential changes in the nasopharyngeal microbiota associated to COVID-19 disease severity, we split the samples into 4 disease severity groups, namely, (1) uninfected control subjects (N = 117); (2) COVID-19 patients with mild symptoms (N = 116); (3) COVID-19 patients with moderate symptoms (N = 97); and (4) COVID-19 patients with severe symptoms (N = 65) (Table S1). Moreover, each of these disease severity groups were divided into 6 age groups covering all stages of life, each divided into females and males to be able to also study possible age- and sex-associated differences (Table S1). Whenever possible, there were 20 samples in each age group and 10 samples in each sex group within them (Table S1). Of the total of 30,535,433 high quality 16S rRNA sequences, 4,427,438 sequences corresponded to the uninfected control subjects (ranging from 10,627 to 256,449 sequences per sample; mean = 37,841; median = 33,134), 7,327,367 sequences corresponded to the COVID-19 patients with mild symptoms (ranging from 8,969 to 223,499; mean = 63,167; median = 53,964), 9,412,508 sequences corresponded to the COVID-19 patients with moderate symptoms (ranging from 43,029 to 299,919; mean = 97,036; median = 82,570), and 9,368,120 sequences corresponded to the COVID-19 patients with severe symptoms (ranging from 71,150 to 330,138; mean = 144,125; median = 137,086). All the previously mentioned 167 families, 329 genera and 671 species were identified in the 4 different disease severity groups established for this study. A simple first analysis of the most abundant taxa at the family, genus, and species levels in each of the COVID-19 disease severity groups (Fig.1a-c), but focusing mainly on the genus level which is the one that can give us more relevant information (as the family level is too general, whereas the taxonomic assignment at the species level may not be fully accurate with the 16S rRNA gene sequencing approach used in this study), revealed some interesting differences (Fig. 1b). Notably, *Dolosigranulum* and *Ralstonia* were among the ten most abundant bacterial genera in uninfected control subjects but were not found in COVID-19 patients with any severity, *Veillonella* was absent only in uninfected controls, and *Rothia* was among the ten most abundant genera only in severe COVID-19 patients while absent in the rest of the severity groups (Fig. 1b). Furthermore, *Acinetobacter* relative abundance was clearly higher in COVID-19 patients compared to uninfected controls (Fig. 1b).

**Figure 1.**
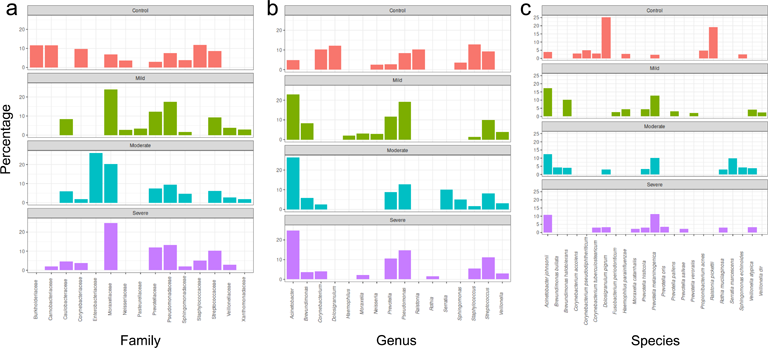
Most abundant taxa in each of the COVID-19 disease severity groups established for this study. Among the 167 families, 329 genera, and 671 species that were identified, all of them present in each of the 4 severity groups, the 10 more abundant taxa at the family (**a**), genus (**b**) and species (**c**) levels in each of the COVID-19 severity groups are shown.

### Clustering patterns of nasopharyngeal samples

With the aim of analyzing how different samples were grouped according to their OTU composition, we applied principal coordinates analysis (PCoA), which is a powerful statistical tool that enables complex multivariate data sets to be visualized in a reduced number of dimensions (23). This allowed us to determine the clustering patterns of samples according to their Bray-Curtis distances, which were calculated based on the relative abundance matrices of the genera across the samples pertinent in each case (Fig. 2). The analysis of similarities (ANOSIM), which is a nonparametric statistical test, was then used to analyse whether there were statistically significant differences among the different age and severity groups included in this study (Fig. 2 and Table S2). The comparison between the different age groups without any stratification by COVID-19 severity showed that, although samples appeared to be mostly intermixed and the different confidence ellipses overlapped each other, there were significant differences in 7 out of the 15 possible comparisons, highlighting the fact that the group containing people over 70 years of age was significantly different to 4 of the other 5 age groups (Fig. 2a and Table S2). Interestingly, the comparison between the COVID-19 disease severity groups without any prior stratification by age revealed that all groups were significantly different from each other (Fig. 2b and Table S2). Then, to study all these differences in more depth, we first stratified the samples by severity and subsequently compared between the different age groups within each severity group (Fig. 2c). A previous work already compared the age groups and analysed the clustering patterns of our control samples according to their Bray-Curtis distances, finding significant differences only between the age groups A1-A4 and A1-A5 (18). When we did the same analysis for the other COVID-19 severity groups, some of the possible comparisons resulted in significant differences in mild and moderate COVID-19 patients (6 out of 15 and 6 out of 10, respectively) (Fig. 2c and Table S2). However, curiously, only there were significant differences between the age groups A3 and A4 in severe COVID-19 patients (Fig. 2c and Table S2). Finally, the samples were stratified first by age and later the COVID-19 severity groups were compared within each age group, finding that, surprisingly, the majority of the possible comparisons were significantly different (24 out of 31) (Fig. 2d and Table S2).

**Figure 2.**
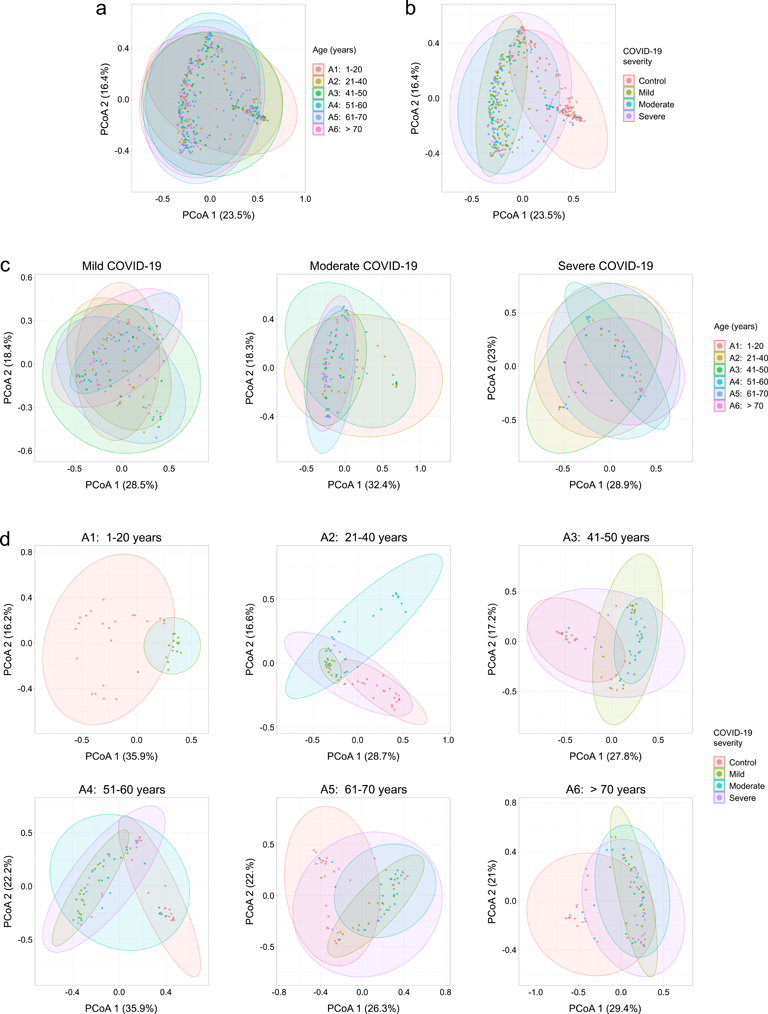
Microbial community composition. Principal coordinates analysis (PCoA) plots of the Bray-Curtis distances which were calculated based on the relative abundance matrices of the genera across the samples pertinent in each case, for the age groups without any prior stratification by COVID-19 disease severity (**a**), for the disease severity groups without any prior stratification by age (**b**), for the age groups within each of the disease severity groups (**c**), and for the disease severity groups within each of the age groups (**d**). In all cases, the data were linearly transformed and visualized in two-dimensional space. Each sample is represented by one dot, colored according to age (**a, c**) or severity (**b, d**). The percentage of the variance of the original data explained by each of the two principal components is indicated in the axis labels. The 90% confidence data ellipses are shown for each age (**a, c**) or severity (**b, d**) group.

### Nasopharyngeal bacterial diversity decreases progressively throughout life in severely ill COVID-19 patients

The fact that previous data on bacterial diversity in the nasopharynx of COVID-19 patients were highly variable and contradictory (14), prompted us to check it as our experimental design avoids many of the limitations that are likely the sources of that variability (14) and, therefore, we could shed light on this issue. Hence, we analysed the alpha diversity, referred to as within-community diversity (24), in the nasopharynx of the different age and severity groups established for this study (Table S1). The Shannon’s diversity index, which measures evenness and richness of communities within a sample, did not show significant changes in bacterial diversity among the different age (Fig. 3a) or severity (Fig. 3b) groups when all the individuals enrolled in this study were included in the analyses without any previous stratification. The only exception was the comparison between COVID-19 patients with mild symptoms and those with moderate ones, since bacterial diversity was significantly lower in the second group (Fig. 3b). Interestingly, when we compared the age groups after having first stratified the individuals by their COVID-19 severity, we found that bacterial diversity progressively decreased throughout life in patients with severe COVID-19, whereas no significant changes between any age groups appeared in COVID-19 patients with mild or moderate symptoms (Fig. 3c). In addition, we have previously demonstrated the absence of any significant changes in alpha diversity in the uninfected control cohort (18). Finally, the comparison between the different COVID-19 severity groups after having first separated the individuals by age, revealed significant changes only between mild and moderate as well as between moderate and severe COVID-19 patients who are in their 20s and 30s (Fig. 3d). To confirm our results, we utilized another index commonly used to measure alpha diversity such as the inverse Simpson’s diversity index, which is an indication of the richness in a community with uniform evenness that would have the same level of diversity. Importantly, results were almost identical to the observed with the Shannon’s diversity index (Fig. S1a-c), including the clear and progressive reduction in bacterial diversity in the nasopharynx of severely ill patients as they age (Fig. S1c).

**Figure 3.**
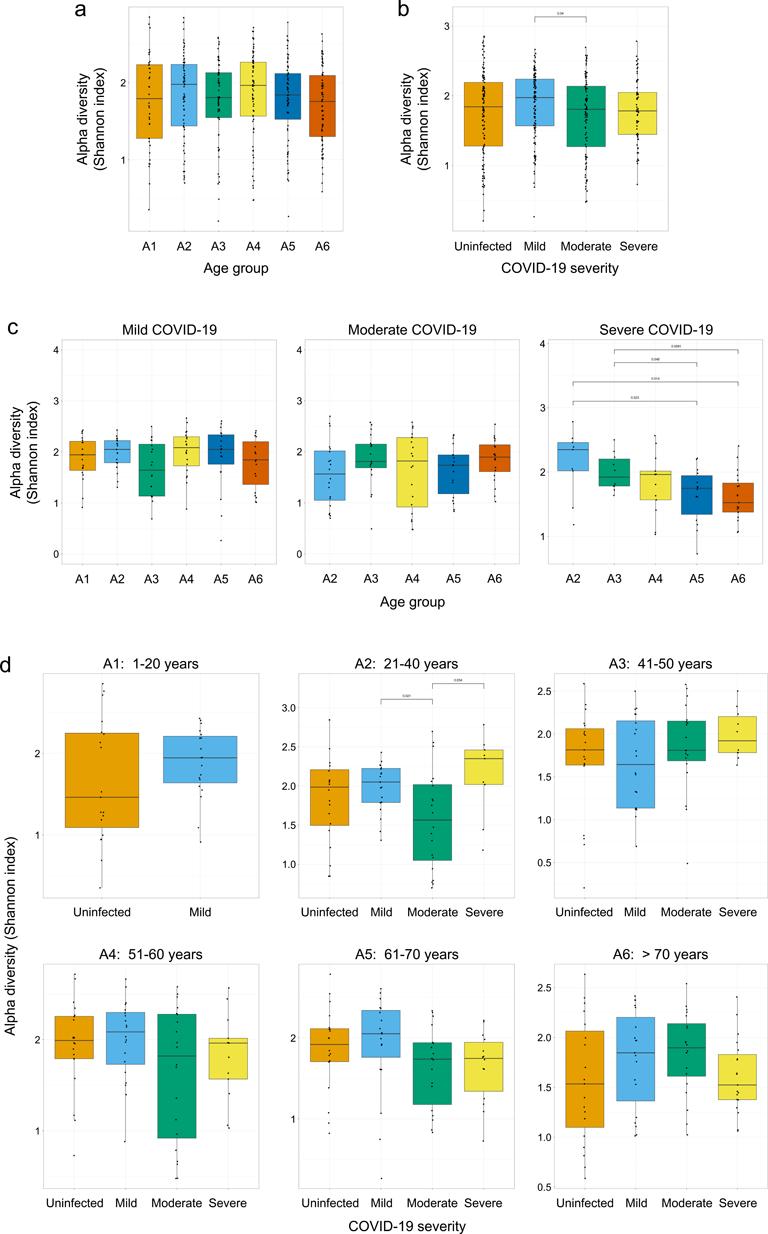
Comparison of alpha diversity parameters across the age and COVID-19 disease severity groups studied. Box-whisker plots showing the alpha diversity Shannon index values for the age groups without any prior stratification by COVID-19 disease severity (**a**), for the disease severity groups without any prior stratification by age (**b**), for the age groups within each of the disease severity groups (**c**), and for the disease severity groups within each of the age groups (**d**). Each sample is represented by one dot. The Wilcoxon signed-rank test was used to determine statistical significance (p-value < 0.05) in the comparisons between the different groups, and, for clarity, only the statistically significant differences and their p-values are showed in the graphs (**a-d**). The age group A1 includes subjects between 1 and 20 years old, A2 between 21 and 40, A3 between 41 and 50, A4 between 51 and 60, A5 between 61 and 70, and A6 includes individuals over 70 years of age (Table S1).

### Age- and severity-associated changes in relative abundance of bacterial taxa in the nasopharynx of COVID-19 patients

We sought to determine the differences in nasopharyngeal taxa abundance among age and severity classes, so we compared the nasopharyngeal microbiota of COVID-19 patients of the different age groups within each of the four disease severity groups established for this study (Fig. 4a), and vice versa (Fig. 4b and Table S1). We considered that focusing our analyses at the genus level would be the most informative since, as previously mentioned, the phylum or family levels are too general, whereas the taxonomic assignment at the species level may not be fully accurate with our 16S rRNA gene sequencing approach. To facilitate the interpretation of a so complex analysis, we selected only the 57 bacterial genera whose relative abundance was higher than 1% in at least one of the age groups and decided to perform heatmap plots to jointly and visually show the results of all the possible comparisons between age groups within each severity group (Fig. 4a), as well as between severity groups within each age group (Fig. 4b). Note that, in some cases, the differences described below were easier to appreciate when the abundance data were represented on a linear scale (Fig. S2), rather than on the more commonly used logarithmic scale that allows to visualize differences when abundance is low (Fig. 4). The comparison between age groups within each of the COVID-19 severity groups showed that, overall, the most abundant bacterial genera shared among all severity groups were *Staphylococcus*, *Dolosigranulum*, *Corynebacterium*, *Streptococcus*, *Moraxella, Prevotella, Acinetobacter, Pseudomonas,* and *Brevundimonas* (Fig. 4a and S2a). However, in the cases of *Streptococcus*, *Prevotella*, *Acinetobacter*, *Pseudomonas*, and *Brevundimonas*, their relative abundances were higher in COVID-19 patients of any severity than in uninfected control individuals (Fig. 4a and S2a). Interestingly, *Sporobacterium*, *Turicella*, and *Cetobacterium* were detected only in uninfected controls of the different age groups whilst they were totally absent in COVID-19 patients independently on their disease severity (Fig. 4a). The cases of *Acinetobacter*, *Pseudomonas*, and *Brevundimonas* were curious as their abundances increased progressively with age in the majority of the severity groups (Fig. 4a and S2a). Notably, the bacterial genera *Spirochaeta* and *Scardovia* were present only in severe COVID-19 patients, being even more interesting the case of *Scardovia* since it was detected exclusively in individuals over 60 years of age (Fig. 4a). As these results suggested a potential utility of *Spirochaeta* and *Scardovia* as biomarkers of COVID-19 disease severity in aged patients, which are the most susceptible to the disease (25), we tried to identify which species of these genera were present in our samples. Thus, while the 16S rRNA gene sequencing approach used in this study did not allow us to identify any *Spirochaeta* species (Fig. 5a), fortunately we were able to determine that the species of *Scardovia* present in our samples was *Scardovia wiggsiae* in more than 99% of the cases (Fig. 5b). Besides confirming all the aforementioned changes in relative abundance, as was logical and expected, the new perspective we had by plotting our relative abundance results as the comparison between the different COVID-19 disease severity groups within each of the age groups, allowed us to observe a higher relative abundance of *Staphylococcus*, *Corynebacterium*, *Streptococcus*, *Prevotella*, *Acinetobacter*, and *Pseudomonas* as the COVID-19 severity increased within most of the age groups (Fig. 4b and S2b).

**Figure 4.**
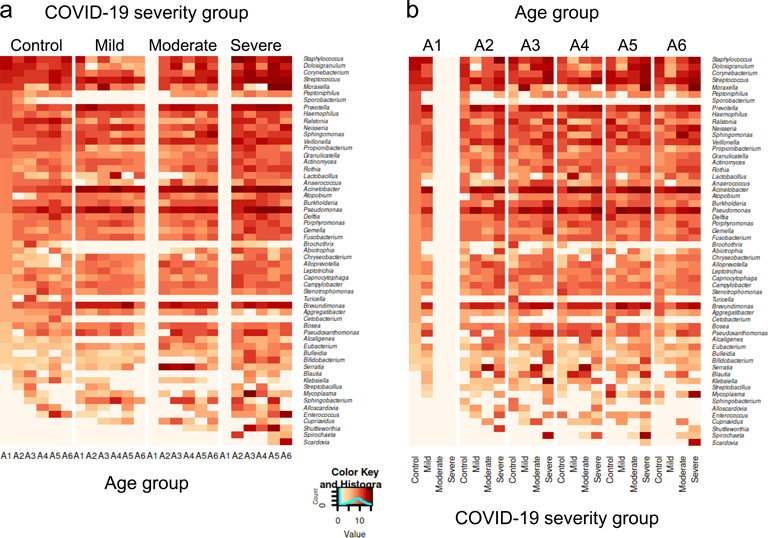
Relative abundance of bacterial genera in the different age and COVID-19 severity groups. Heatmaps showing the abundance for each of the age and severity groups established for this study of the 57 bacterial genera whose relative abundance is above 1% in at least one of the age groups. Data are showed in two different ways to facilitate the visualization and interpretation of such a complex data set: divided first by severity group and then by age group within each severity group (**a**), and divided first by age group and then by severity group within each age group (**b**). Data are shown on a logarithmic scale to facilitate the visualization of differences when abundance is low. Bacterial genera are arranged in decreasing order of abundance considering the first column on the left, which corresponds to the uninfected control subjects of the age group A1.

**Figure 5.**
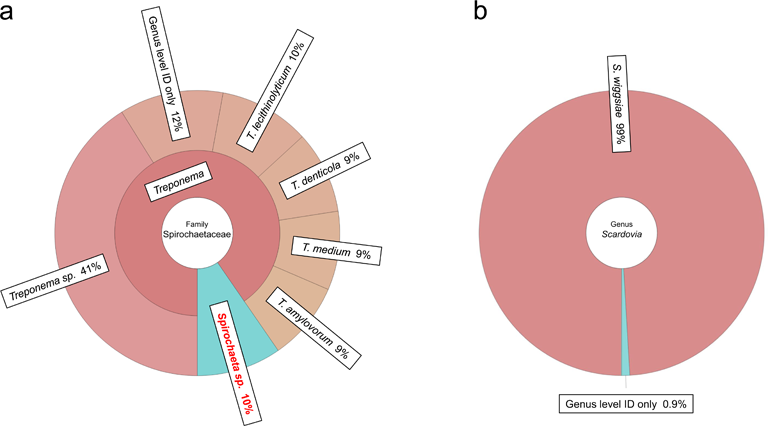
Bacterial community composition of the family Spirochaetaceae and the genus *Scardovia* in severely ill COVID-19 patients over 70 years of age. **a** Krona chart of the bacterial community composition of the family Spirochaetaceae in severe COVID-19 patients over 70 years of age, showing that our 16S rRNA gene sequencing approach was not able to go beyond the genus level in the case of the bacterial genus *Spirochaeta*. **b** Krona chart of the bacterial community composition of the genus *Scardovia* in severe COVID-19 patients over 70 years of age, showing that the bacteria from this genus was *Scardovia wiggsiae* in more than 99% of cases.

Finally, aiming to identify any statistically significant age-associated changes in the nasopharyngeal microbiota of COVID-19 patients, we focused on the bacterial genera whose relative abundances were significantly different between the distinct age groups within each of the severity groups (Fig. 6). We had already been reported in a previous study with the uninfected control cohort that there were statistically significant differences in relative abundance between the distinct age groups in the 11 bacterial genera *Acinetobacter*, *Brevundimonas*, *Dolosigranulum*, *Finegoldia*, *Haemophilus*, *Leptotrichia*, *Moraxella*, *Peptoniphilus*, *Pseudomonas*, *Rothia*, and *Staphylococcus* (18). Curiously, our results showed that the number of bacterial genera which presented statistically significant differences in relative abundance between the age groups clearly decreased as the COVID-19 disease severity increased, with 23 genera presenting such differences in COVID-19 patients with mild symptoms (Fig. 6a and Table S3), 15 genera in patients with moderate symptoms (Fig. 6b and Table S3), and only 4 genera in individuals suffering from severe COVID-19 (Fig. 6c and Table S3). Differences between age groups were statistically significant in 74 cases in the 23 bacterial genera that presented this type of differences in mild COVID-19 patients, in 52 cases in the 15 genera that presented them in moderate COVID-19 patients, and in 13 cases in the 4 genera that presented them in severely ill COVID-19 patients (Table S3). Curiously, the age group A6 was the one that presented statistically significant differences with respect to other age groups in more cases in mild and moderate COVID-19 patients, but not in patients with severe symptoms (Table S3).

**Figure 6.**
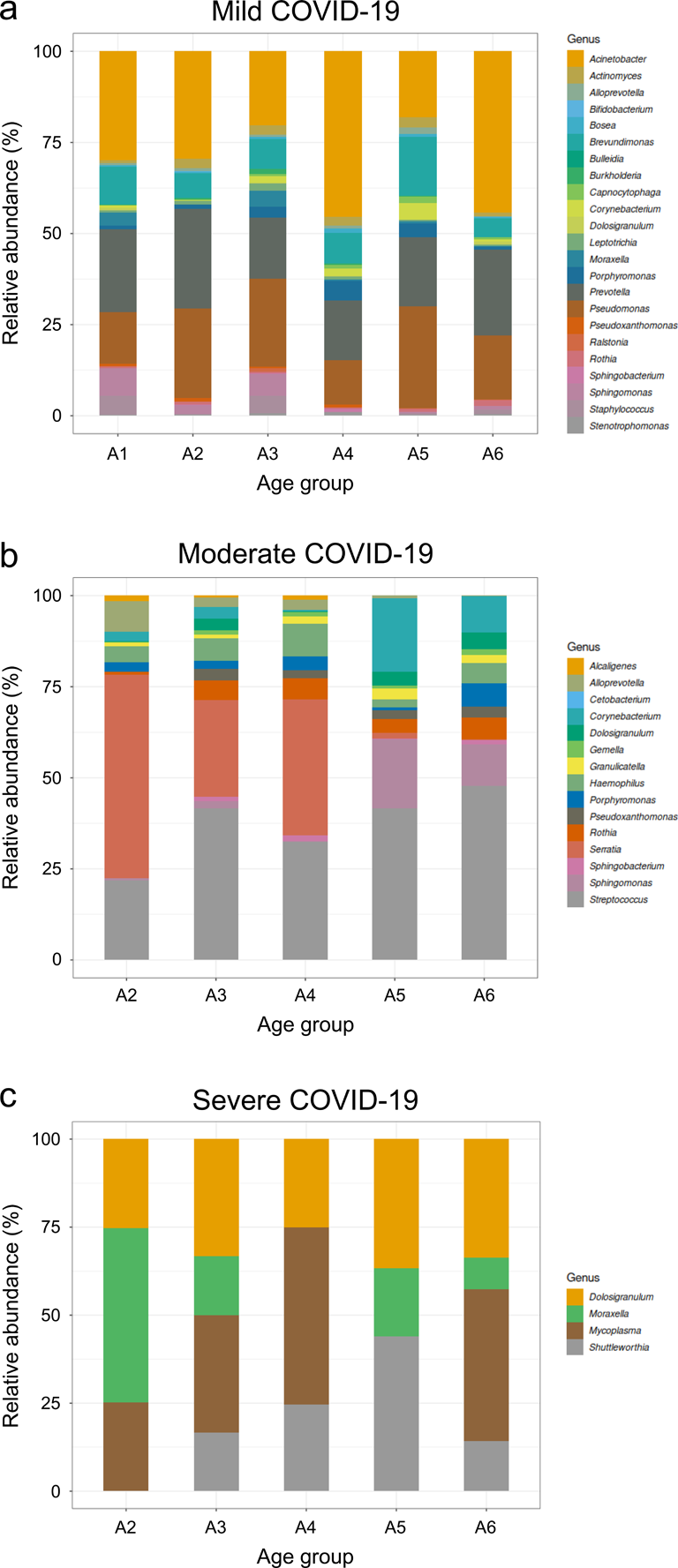
Taxonomic composition of the bacterial genera which show significant differences between age groups in mild, moderate, and severe COVID-19 patients. Stacked bar charts showing the relative abundance (%) of the bacterial genera that present statistically significant differences between age groups in mild (**a**), moderate (**b**), and severe (**c**) COVID-19 patients. Bacterial genera are arranged in alphabetical order.

## DISCUSSION

After characterizing the healthy human nasopharyngeal microbiota (18), we sought to study any possible links between alterations of this microbiota and SARS-CoV-2 infection, with special attention to the COVID-19 clinical outcome, as this is a crucial question from a biomedical and clinical point of view that remains unresolved. But the problem is not the lack of studies on this, since dozens of works have already addressed this topic, but rather that their results are extremely variable and contradictory (14). In a recent previous work, we identified and discussed in depth the potential sources of such high variability study by study, highlighting the low sample sizes, the heterogeneity of the enrolled subjects, the different sequencing technologies used, or the lack of standardization of the criteria utilized to stratify individuals, among others (14). Thus, in the present study we have tried to avoid the mentioned sources of variability, as far as possible, aiming to obtain soundness data on any significant sex-, age-, or disease severity-associated changes in the nasopharyngeal microbiota of COVID-19 patients. Regarding sex-associated changes, no significant differences in any of the parameters analysed in this study were found at all, including within any of the age or COVID-19 disease severity groups investigated. This is consistent with the findings of a previous study using our uninfected control cohort to characterize the nasopharyngeal microbiota of healthy subjects, that revealed the absence of any relevant differences between males and females (18), and confirms that nasopharyngeal microbiota does not behave as that of other anatomical areas such as the gut, where there are significant sex-associated differences in diversity and taxonomic composition, probably due to the distinct levels of sex hormones (12, 26, 27). Therefore, analyses by sex are not showed in this work given their total irrelevance and to simplify the analysis and interpretation of the age- and disease severity-associated differences.

Our initial PCoA analyses to determine the clustering patterns of the nasopharyngeal samples revealed that, without any prior stratification by COVID-19 disease severity, the age group containing people over 70 years of age was the most different from the rest of age groups. This was the first hint that the results of the oldest people were going to be some of the most interesting findings throughout this study, as will be seen below. After this promising beginning, we decided to analyse alpha diversity. It had previously been observed that alpha diversity, which summarizes the distribution of species abundances in a given sample into a single number that depends on species richness and evenness, and is a central topic in microbiome data analysis (24), was not significantly different in any case when comparing between age groups in the uninfected cohort used as a control in this study (18). Similarly, analysing bacterial species richness in the nasopharynx of subjects of the different age and COVID-19 disease severity groups established for this study, revealed no age- or severity-associated relevant differences in alpha diversity, with the only striking exception of COVID-19 patients with severe symptoms, whose alpha diversity decreased progressively as their age increased. This was confirmed by using two of the most reliable and commonly used alpha diversity indexes, such as the Shannon’s diversity index and the inverse Simpson’s diversity index, which resulted in almost identical results. The general lack of differences in alpha diversity found in this work is consistent with the results of many previous studies that observed the same when comparing the nasopharyngeal microbiota of COVID-19 patients with that of uninfected controls (28–34), whereas, in accordance with the variability prevailing in this field until now, other studies had reported that alpha diversity decreased in COVID-19 patients compared to uninfected controls (35, 36), that it decreased in the most severe cases of COVID-19 compared to milder cases (37–40), and even that it increased in SARS-CoV-2 infected pregnant women compared to uninfected (41). Unfortunately, the already mentioned limitations present in all these previous studies (14), together with the fact that their experimental designs did not allow the analysis of the nasopharyngeal microbiota of COVID-19 patients with different disease severities at all stages of their lives, prevented them from finding the progressive alpha diversity reduction in severe COVID-19 patients uncovered in this work. Curiously, similar reductions in alpha diversity with aging had been observed before in the gut of healthy subjects (42), but in that case the differences are probably due to aging-associated factors that affect the gut microbiota in a more intense way compared to the nasopharyngeal microbiota, such as the increase of coliform numbers and changes in diet (43). In summary, our analyses have revealed for the first time, to our knowledge, that alpha diversity progressively decreases with aging but only in patients with severe COVID-19, strongly suggesting an association between this reduced bacterial diversity and the fact that aged individuals are more susceptible to COVID-19 and present more severe forms of the disease (6).

The use of heatmaps, that represent the magnitude of individual values within a dataset as a colour, allowed us to analyse our complex taxa abundance results at the genus level in a simpler and more visual manner. Cases in which a bacterial genus is present or absent exclusively in a certain COVID-19 disease severity group are particularly relevant from a biomedical point of view, since the presence/absence of such microorganism could potentially serve as a biomarker of disease severity. This is exactly what we observed for the bacterial genera *Sporobacterium*, *Turicella* and *Cetobacterium*, which were abundant in uninfected individuals while totally absent in COVID-19 patients independently on their disease severity. Even more striking was the finding that *Spirochaeta* and *Scardovia* were present only in COVID-19 patients with severe symptoms, being particularly relevant the case of *Scardovia* as it appeared exclusively in subjects over 60 years of age. The 16S rRNA gene sequencing approach used in this study, which is by far the most common technique to study microbiota in clinical samples, has the limitation that its taxonomic resolution often does not allow identification beyond the genus level (44), as we found in the case of *Spirochaeta*. However, fortunately, our analyses were able to determine that the bacteria of the genus *Scardovia* identified in severe COVID-19 patients over 60 years of age were of the species *S. wiggsiae* in more than 99% of the cases. *S. wiggsiae*, a gram-positive, anaerobic, nonspore forming, and nonmotile bacilli removed from the genus *Bifidobacterium* in 2002 due to difference in its genome sequence (45), was classified as part of the human oral microbiome (46) and later identified as a predominant caries pathogen, even in the absence of *Streptococcus mutans* (47, 48). Furthermore, our novel finding that *Scardovia* was present only in the nasopharynx of severe COVID-19 patients over 60 years of age was consistent with a previous study that analysed the nasal/oropharyngeal microbiota of COVID-19 patients with different disease severities, where *Scardovia* was detected only in intensive care unit patients, albeit unfortunately those patients were not stratified by age (49). Therefore, all these data together suggest that while bacteria from the genus *Scardovia* are present in the oral microbiota (46), they are also capable of colonizing other close ecological niches such as the nose, oropharynx, and nasopharynx in severely ill COVID-19 patients (49). Although it seems counterintuitive, it is well known that nearby anatomical areas closely related in terms of structure and function can present different microbiotas and even niche-specific bacteria, as previously demonstrated, for example, for the cases of nasopharynx and nose that are adjacent (50). In the present case, thinking of the possible changes in the nasopharynx of severely ill COVID-19 patients that allow *Scardovia* to colonize new anatomical areas of their upper respiratory tract, such alterations may be caused by the replication of the SARS-CoV-2 virus itself or by the inflammatory processes in response to the infection that disrupt the physical-chemical barriers. Undoubtedly, this issue will deserve further research. Note that even though the intubation procedures can also alter the nasopharynx in severe patients, this is not applicable to the present study as our samples were collected in early stages of the disease. In addition, it was particularly concerning, from a clinical perspective, that the opportunistic pathogen bacterial genera *Staphylococcus*, *Corynebacterium*, *Streptococcus*, *Prevotella*, *Acinetobacter*, and *Pseudomonas* were overrepresented in the nasopharynx of severely ill COVID-19 patients. An important question here is whether the previously mentioned changes in the nasopharynx as consequence of a severe COVID-19 disease allow a greater proliferation of these opportunistic microorganisms in the nasopharynx, or whether, on the contrary, a greater previous abundance of these bacterial genera in the nasopharynx predisposes subjects to suffer more serious forms of the COVID-19 disease. Since, as already mentioned, our samples were collected in early stages of the disease, the second hypothesis is probably the correct one.

Our taxa abundance analyses at the genus level detected statistically significant relative abundance differences between the distinct age groups for 23 bacterial genera in COVID-19 patients with mild symptoms, for 15 genera in patients with moderate disease severity, and only for 4 genera in severely ill patients. The reason why the number of bacterial genera presenting significant differences between age groups decreased as the COVID-19 disease severity increased is enigmatic, but we can hypothesize that the previously mentioned changes induced by the COVID-19 disease alter the conditions of the nasopharynx as an ecological niche to such an extent at any age, that it determines which bacterial genera can live there independently of the age of the patients. In other words, the significant relative abundance differences between age groups observed in COVID-19 patients with mild or moderate severities, could be counteracted in severe patients by a more powerful factor such as the alterations in the nasopharynx caused by the COVID-19 disease, proportionally to its severity. This hypothesis is supported by the fact that 52.7% (39 out of 74) of the relative abundance statistically significant differences in mild COVID-19 patients, 67.3% (35 out of 52) in moderate patients, whereas only 23.1% (3 out of 13) in severe patients, were between the age group A6 and other age groups. Thus, while A6 clearly is the most different group from the other age groups in mild and moderate COVID-19 patients, this dynamic is broken in severe patients where probably the alterations as consequence of the disease are the predominant factor.

As previously mentioned, this study was designed trying to avoid, as much as possible, the limitations that we identified in a recent work (14) as the greatest sources of data variability on this field. Good examples of this are our high sample size, our stratification of subjects by age in groups that cover the entire life, or how our samples were collected from a very short period of time that comprised a single SARS-CoV-2 infection wave at an early point in the pandemic, thus avoiding the risk of recruiting patients infected by different SARS-CoV-2 variants that are already known to elicit different immune responses that could alter the nasopharyngeal microbiota differentially (51). Nevertheless, our study still has several limitations. This was an observational, retrospective study, and collection of data was not standardized in advance. Even though it is the most common technique to study microbiota in clinical simples, the 16S rRNA gene sequencing approach to study the microbiota could introduce bias in the obtained data because this method does not allow the study of the whole microbiome, but only the genera amplified by PCR. Moreover, it was not possible to obtain serial samples. Furthermore, the sex groups within each age group are small, so the study may have been underpowered to detect certain associations. Finally, we could not access any sociodemographic, environmental, lifestyle, or medical information of subjects enrolled in this study, which would have been helpful to better understand the characteristics of the cohort.

Notably, some of the most clear and interesting differences found in this study were those that affected the elderly, who are the most susceptible to developing serious forms of the COVID-19 disease (25). Thus, they were (1) the more different age group compared to the other age groups, without any prior stratification by COVID-19 disease severity, according to our PCoA analyses; (2) the age group that presented a greater reduction in alpha diversity in severely ill COVID-19 patients; (3) the age group involved in more comparisons between age groups that resulted in statistically significant differences, and (4) only severe COVID-19 patients over 60 years of age presented the bacteria *S. wiggsiae*-potentially useful as a severity biomarker in these individuals-in their nasopharynx. In addition, the relative abundances of opportunistic bacterial pathogens such as *Moraxella* and *Acinetobacter* were increased in aged severe COVID-19 patients compared with younger patients with the same disease severity. Therefore, we can hypothesize that there may be some correlation between the increased susceptibility of aged subjects to COVID-19 (25), and their nasopharyngeal taxonomic composition. Hence, future metagenomic studies collecting samples at different time points will be paramount to elucidate when the mentioned changes in the nasopharyngeal microbiota of aged individuals occur, aiming to determine whether such changes are cause or consequence of the COVID-19 disease and, therefore, if they could be useful as prognosis or severity biomarkers in the elderly. Moreover, this could also be useful to stratify COVID-19 aged patients, and paves the way for new therapeutic avenues such as nasopharyngeal microbiota manipulation, which is an approach that has already been successfully exploited in different medical fields, from cancer to metabolic disorders and viral infection (15–17, 52).

## Data Availability

Raw sequencing data of all 16S rRNA sequences and metadata are available at the open access repository Figshare under the accession numbers 10.6084/m9.figshare.24504259 and 10.6084/m9.figshare.24504262, respectively.

## Acknowledgements

We thank the staff of the Genomics facility of IMIB Pascual Parrilla for 16S rRNA sequencing, and the staff of the Microbiology Service at HCUVA for sample collection and processing.

## Author contributions

The authors offer the following declarations about their contributions: Conceived and designed the experiments: VM, MLC and SC. Performed the experiments: FPS, SDT, CAS and SC. Analysed the data: FPS, SDT, CAS, VM, MLC and SC. Provided patients’ samples: AMD. Writing-original draft: SC. Writing-review & editing: VM and MLC. All authors have read and agreed to the published version of the manuscript.

## Institutional review board statement

All procedures in this work were carried out following the principles expressed in the Declaration of Helsinki, as well as in all the other applicable international, national, and/or institutional guidelines for the use of samples and data, and have been approved by the Comité de Ética de la Investigación (CEIm) at Hospital Clínico Universitario Virgen de la Arrixaca (protocol number 2020-10-12-HCUVA—Effects of aging in the susceptibility to SARS-CoV-2).

## Informed consent statement

Nasopharyngeal swabs were collected for diagnosis of SARS-CoV-2 infection before this study was conceived, without the need of any informed consent as the collection procedure was non-invasive and risk-free. However, when the COVID-19 pandemic spread out of control, samples were kept at the Microbiology Service instead of destroyed after diagnosis as it was considered that they might be extremely relevant for research. This, together with the facts that (i) the retrospective use of these samples did not affect donor health or treatment at all, (ii) all data has been treated anonymously, and (iii) movement was limited due to the exceptional circumstances of the pandemic meant that it was not possible to obtain informed consents for the use of these samples in research. Moreover, none of the subjects expressly objected to their samples being used for research.

---

**Figure S1.**
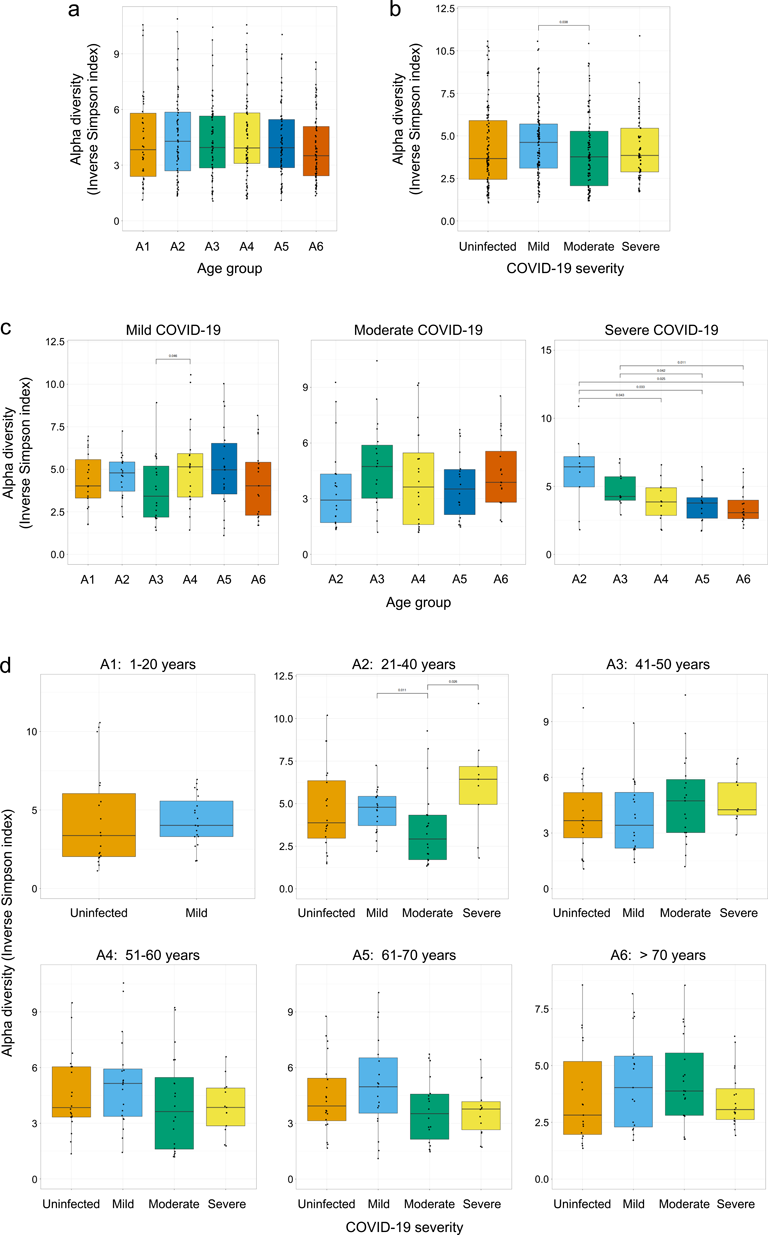
Comparison of alpha diversity parameters across the age and COVID-19 disease severity groups studied. Box-whisker plots showing the alpha diversity inverse Simpson index values for the age groups without any prior stratification by COVID-19 disease severity (**a**), for the disease severity groups without any prior stratification by age (**b**), for the age groups within each of the disease severity groups (**c**), and for the disease severity groups within each of the age groups (**d**). Each sample is represented by one dot. The Wilcoxon signed-rank test was used to determine statistical significance (p-value < 0.05) in the comparisons between the different groups, and, for clarity, only the statistically significant differences and their p-values are showed in the graphs (**a-d**). The age group A1 includes subjects between 1 and 20 years old, A2 between 21 and 40, A3 between 41 and 50, A4 between 51 and 60, A5 between 61 and 70, and A6 includes individuals over 70 years of age (Table S1).

**Figure S2.**
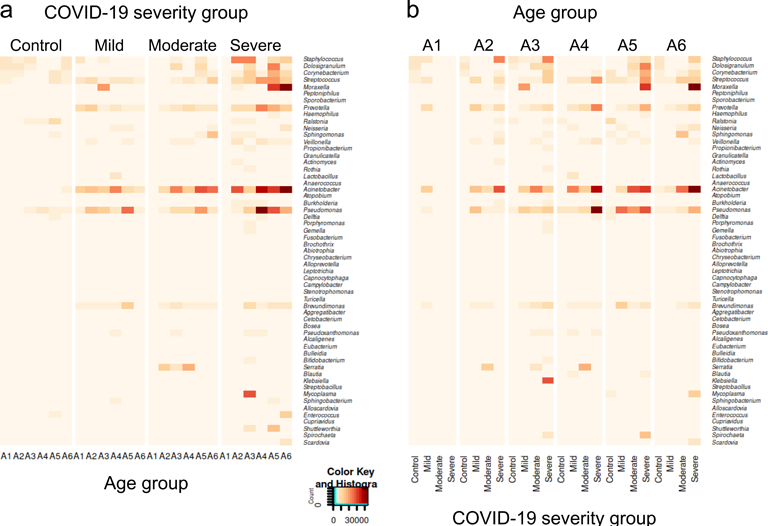
Relative abundance of bacterial genera in the different age and COVID-19 severity groups. Heatmaps showing the abundance for each of the age and severity groups established for this study of the 57 bacterial genera whose relative abundance is above 1% in at least one of the age groups. Data are showed in two different ways to facilitate the visualization and interpretation of such a complex data set: divided first by severity group and then by age group within each severity group (**a**), and divided first by age group and then by severity group within each age group (**b**). Data are shown on a linear scale to facilitate the visualization of differences when abundance is high. Bacterial genera are arranged in decreasing order of abundance considering the first column on the left, which corresponds to the uninfected control subjects of the age group A1.

**Table S1.** Nasopharyngeal exudate samples collected for this study.

| Age group | Sex | Uninfected control individuals | COVID-19 patients |  |  |
| --- | --- | --- | --- | --- | --- |
|  |  |  | Mild severity | Moderate severity | Severe severity |
| A1: 1-20 years | *Female* | 10 | 9 | 0 | 0 |
|  | *Male* | 9 | 10 | 0 | 0 |
| A2: 21-40 years | *Female* | 10 | 10 | 10 | 5 |
|  | *Male* | 10 | 9 | 10 | 4 |
| A3: 41-50 years | *Female* | 9 | 10 | 10 | 0 |
|  | *Male* | 10 | 10 | 9 | 10 |
| A4: 51-60 years | *Female* | 10 | 10 | 10 | 5 |
|  | *Male* | 10 | 10 | 10 | 8 |
| A5: 61-70 years | *Female* | 10 | 10 | 8 | 7 |
|  | *Male* | 10 | 9 | 10 | 7 |
| A6: >70 years | *Female* | 9 | 10 | 10 | 9 |
|  | *Male* | 10 | 9 | 10 | 10 |
| Total | *Female* | 58 | 59 | 48 | 26 |
|  | *Male* | 59 | 57 | 49 | 39 |

**Table S2.**
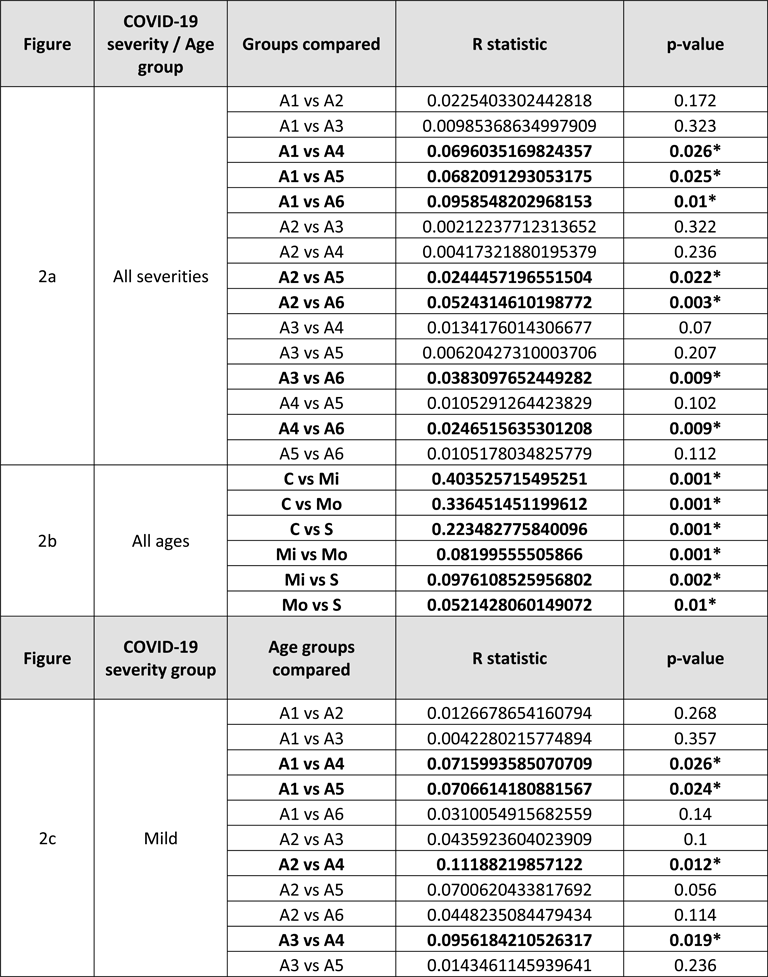
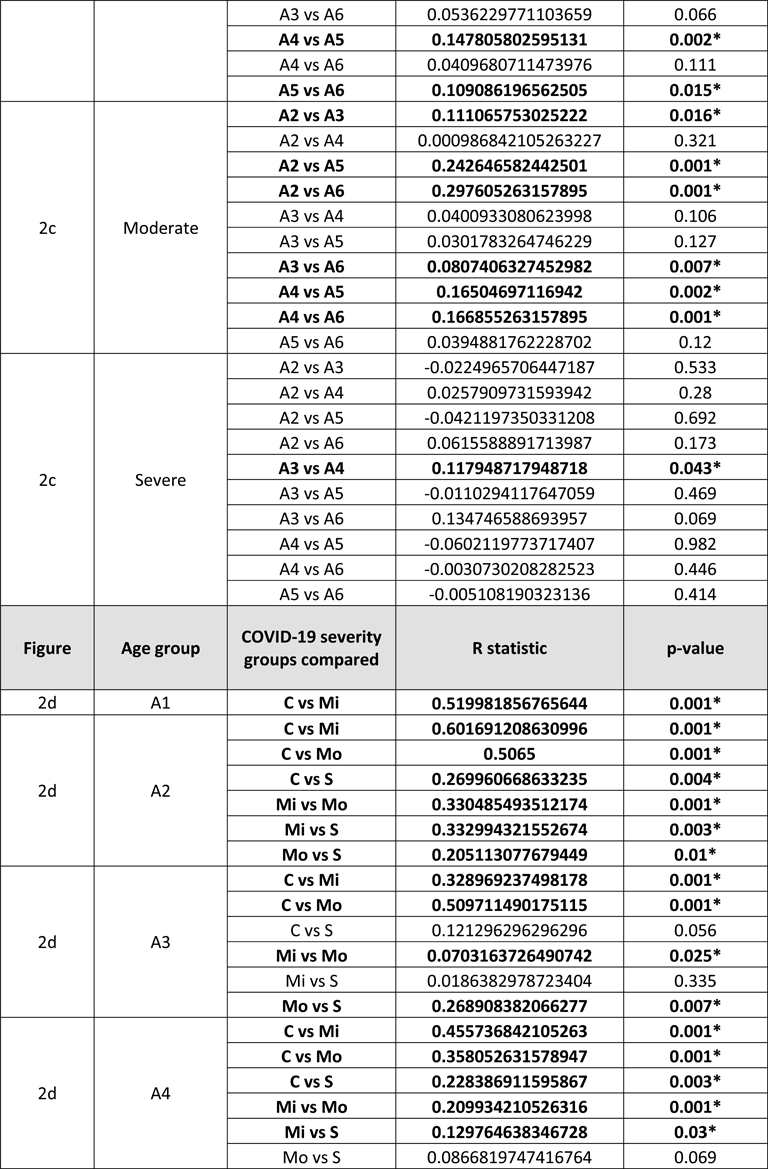
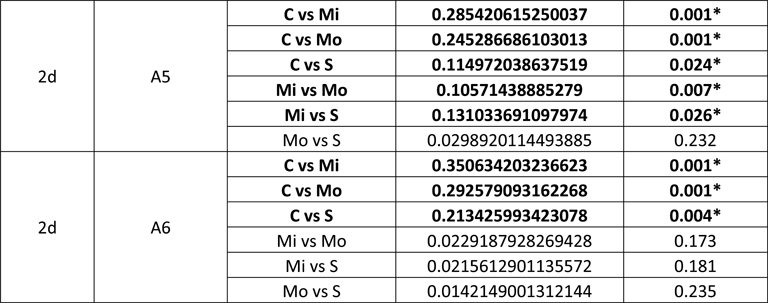
Statistical analysis of the PCoA results for all the possible comparisons between age or COVID-19 disease severity groups as indicated, according to the nonparametric statistical test ANOSIM. The statistically significant differences (p-value < 0.05) are highlighted in bold letter, and their p-values are marked with an asterisk. C, control; Mi, mild; Mo, moderate; S, severe.

**Table S3.** Summary of the statistical analysis of the relative abundance differences between the age groups established in this study for mild, moderate, and severe COVID-19 patients, as indicated. Only the statistically significant differences (adjusted p-value < 0.05) are shown.

| MILD COVID-19 SEVERITY |  |  |  |  |  |  |  |
| --- | --- | --- | --- | --- | --- | --- | --- |
| Genus | baseMean | log2FoldChange | lfcSE | stat | p-value | Adjusted p-value | Age groups compared |
| *Acinetobacter* | 15127,961 | 1,753 | 0,578 | 3,035 | 0,002 | 0,048 | A4_A5 |
| *Actinomyces* | 398,074 | 2,29 | 0,853 | 2,683 | 0,007 | 0,036 | A2_A6 |
| *Alloprevotella* | 187,097 | 3,231 | 1,059 | 3,053 | 0,002 | 0,015 | A2_A6 |
|  |  | 3,496 | 1,045 | 3,344 | 0,001 | 0,006 | A4_A6 |
|  |  | 2,702 | 1,059 | 2,552 | 0,011 | 0,048 | A5_A6 |
| *Bifidobacterium* | 16,556 | 4,71 | 1,426 | 3,303 | 0,001 | 0,013 | A1_A5 |
|  |  | 3,934 | 1,388 | 2,834 | 0,005 | 0,031 | A2_A4 |
|  |  | 5,55 | 1,424 | 3,896 | 0 | 0,001 | A2_A5 |
|  |  | -4,963 | 1,427 | -3,479 | 0,001 | 0,004 | A5_A6 |
| *Bosea* | 401,565 | -2,873 | 0,997 | -2,883 | 0,004 | 0,026 | A1_A4 |
|  |  | -3,084 | 0,997 | -3,095 | 0,002 | 0,016 | A2_A4 |
|  |  | 3,343 | 0,997 | 3,353 | 0,001 | 0,006 | A4_A6 |
| *Brevundimonas* | 5060,261 | 2,3 | 0,639 | 3,6 | 0 | 0,003 | A3_A6 |
|  |  | 2,645 | 0,647 | 4,088 | 0 | 0 | A5_A6 |
| *Bulleidia* | 6,264 | 5,172 | 1,636 | 3,16 | 0,002 | 0,016 | A1_A4 |
|  |  | 4,922 | 1,648 | 2,987 | 0,003 | 0,023 | A1_A5 |
|  |  | 5,175 | 1,675 | 3,089 | 0,002 | 0,013 | A1_A6 |
|  |  | 4,262 | 1,676 | 2,543 | 0,011 | 0,045 | A2_A6 |
| *Burkholderia* | 62,377 | 20,588 | 2,715 | 7,582 | 0 | 0 | A1_A6 |
|  |  | 21,654 | 2,711 | 7,987 | 0 | 0 | A2_A6 |
|  |  | 26,799 | 2,676 | 10,016 | 0 | 0 | A3_A6 |
|  |  | 24,401 | 2,676 | 9,119 | 0 | 0 | A4_A6 |
|  |  | 21,061 | 2,714 | 7,761 | 0 | 0 | A5_A6 |
| *Capnocytophaga* | 86,147 | -4,642 | 1,551 | -2,992 | 0,003 | 0,023 | A1_A5 |
| *Corynebacterium* | 92,239 | -4,113 | 1,184 | -3,474 | 0,001 | 0,005 | A2_A4 |
|  |  | -4,238 | 1,199 | -3,535 | 0 | 0,004 | A2_A5 |
|  |  | -3,04 | 1,2 | -2,534 | 0,011 | 0,045 | A2_A6 |
| *Dolosigranulum* | 51,792 | 23,977 | 1,997 | 12,005 | 0 | 0 | A1_A2 |
|  |  | -21,122 | 1,984 | -10,645 | 0 | 0 | A2_A3 |
|  |  | -25,587 | 1,972 | -12,974 | 0 | 0 | A2_A4 |
|  |  | -24,081 | 1,997 | -12,057 | 0 | 0 | A2_A5 |
|  |  | -27,439 | 1,996 | -13,75 | 0 | 0 | A2_A6 |
|  |  | -6,317 | 1,929 | -3,275 | 0,001 | 0,008 | A3_A6 |
| *Leptotrichia* | 224,389 | 2,873 | 1,148 | 2,503 | 0,012 | 0,045 | A2_A6 |
| *Moraxella* | 130,937 | 6,35 | 1,538 | 4,13 | 0 | 0,001 | A1_A2 |
|  |  | 4,618 | 1,515 | 3,047 | 0,002 | 0,018 | A1_A4 |
|  |  | 5,548 | 1,539 | 3,606 | 0 | 0,002 | A1_A6 |
| *Porphyromonas* | 475,245 | 2,47 | 0,712 | 3,468 | 0,001 | 0,006 | A4_A6 |
|  |  | 1,911 | 0,721 | 2,649 | 0,008 | 0,044 | A5_A6 |
| *Prevotella* | 5394,065 | 1,457 | 0,524 | 2,781 | 0,005 | 0,031 | A2_A6 |
| *Pseudomonas* | 14997,008 | -2,841 | 0,691 | -4,112 | 0 | 0,002 | A1_A3 |
|  |  | -2,026 | 0,7 | -2,896 | 0,004 | 0,025 | A1_A5 |
|  |  | 2,529 | 0,682 | 3,709 | 0 | 0,008 | A3_A4 |
|  |  | 2,65 | 0,691 | 3,835 | 0 | 0,002 | A3_A6 |
|  |  | 1,835 | 0,7 | 2,623 | 0,009 | 0,044 | A5_A6 |
| *Pseudoxanthomonas* | 348,769 | 7,303 | 2,4 | 3,043 | 0,002 | 0,015 | A4_A6 |
| *Ralstonia* | 8,671 | 22,523 | 2,143 | 10,51 | 0 | 0 | A1_A6 |
|  |  | 20,518 | 2,147 | 9,555 | 0 | 0 | A2_A6 |
|  |  | 23,15 | 2,117 | 10,937 | 0 | 0 | A3_A6 |
|  |  | 19,258 | 2,133 | 9,03 | 0 | 0 | A4_A6 |
|  |  | 20,319 | 2,15 | 9,452 | 0 | 0 | A5_A6 |
| *Rothia* | 147,817 | -3,176 | 1,094 | -2,902 | 0,004 | 0,025 | A3_A6 |
| *Sphingobacterium* | 14,4 | -15,84 | 4,853 | -3,264 | 0,001 | 0,022 | A1_A3 |
|  |  | -24,17 | 4,834 | -5 | 0 | 0 | A1_A4 |
|  |  | -20,282 | 4,9 | -4,14 | 0 | 0 | A1_A6 |
|  |  | -25,575 | 4,853 | -5,27 | 0 | 0 | A2_A3 |
|  |  | -33,905 | 4,834 | -7,013 | 0 | 0 | A2_A4 |
|  |  | -30,016 | 4,9 | -6,126 | 0 | 0 | A2_A6 |
|  |  | 25,192 | 4,853 | 5,191 | 0 | 0 | A3_A5 |
|  |  | 33,522 | 4,834 | 6,934 | 0 | 0 | A4_A5 |
|  |  | -29,633 | 4,9 | -6,048 | 0 | 0 | A5_A6 |
| *Sphingomonas* | 975,21 | 5,627 | 1,114 | 5,052 | 0 | 0 | A1_A4 |
|  |  | 5,147 | 1,128 | 4,564 | 0 | 0 | A1_A5 |
|  |  | 4,115 | 1,128 | 3,649 | 0 | 0,002 | A1_A6 |
|  |  | 5,265 | 1,114 | 4,726 | 0 | 0 | A2_A4 |
|  |  | 4,785 | 1,128 | 4,243 | 0 | 0 | A2_A5 |
|  |  | 3,753 | 1,128 | 3,328 | 0,001 | 0,007 | A2_A6 |
| *Staphylococcus* | 104,676 | 3,362 | 1,12 | 3,002 | 0,003 | 0,036 | A1_A2 |
|  |  | 3,957 | 1,107 | 3,574 | 0 | 0,005 | A1_A4 |

|  |  | 5,28 | 1,127 | 4,687 | 0 | 0 | A1_A5 |
| --- | --- | --- | --- | --- | --- | --- | --- |
|  |  | 4,465 | 1,113 | 4,013 | 0 | 0,001 | A3_A5 |
|  |  | -3,167 | 1,128 | -2,809 | 0,005 | 0,033 | A5_A6 |
| *Stenotrophomonas* | 291,969 | -2,472 | 0,936 | -2,641 | 0,008 | 0,047 | A1_A4 |
|  |  | 2,538 | 0,936 | 2,71 | 0,007 | 0,037 | A4_A6 |
| **MODERATE COVID-19 SEVERITY** |  |  |  |  |  |  |  |
| **Genus** | **baseMean** | **log2FoldChange** | **lfcSE** | **stat** | **p-value** | **Adjusted p-value** | **Age groups compared** |
| *Alcaligenes* | 395,263 | 7,3 | 1,439 | 5,074 | 0 | 0 | A2_A5 |
|  |  | 8,774 | 1,417 | 6,191 | 0 | 0 | A2_A6 |
|  |  | 4,515 | 1,457 | 3,099 | 0,002 | 0,028 | A3_A5 |
|  |  | 5,989 | 1,436 | 4,172 | 0 | 0 | A3_A6 |
|  |  | 8,715 | 1,439 | 6,058 | 0 | 0 | A4_A5 |
|  |  | 10,189 | 1,417 | 7,19 | 0 | 0 | A4_A6 |
| *Alloprevotella* | 242,803 | 6,103 | 1,768 | 3,451 | 0,001 | 0,002 | A2_A6 |
|  |  | 5,379 | 1,791 | 3,003 | 0,003 | 0,013 | A3_A6 |
|  |  | 5,358 | 1,768 | 3,03 | 0,002 | 0,018 | A4_A6 |
| *Cetobacterium* | 0,934 | -20,683 | 4,518 | -4,577 | 0 | 0 | A2_A6 |
|  |  | -27,986 | 4,578 | -6,113 | 0 | 0 | A3_A6 |
|  |  | -28,371 | 4,519 | -6,279 | 0 | 0 | A4_A6 |
|  |  | -27,685 | 4,644 | -5,962 | 0 | 0 | A5_A6 |
| *Corynebacterium* | 846,952 | -3,07 | 1,089 | -2,819 | 0,005 | 0,045 | A3_A5 |
|  |  | -3,329 | 1,061 | -3,138 | 0,002 | 0,01 | A3_A6 |
|  |  | -3,004 | 1,047 | -2,87 | 0,004 | 0,025 | A4_A6 |
| *Dolosigranulum* | 126,882 | -6,144 | 2,214 | -2,775 | 0,006 | 0,018 | A2_A6 |
|  |  | -6,454 | 2,31 | -2,794 | 0,005 | 0,045 | A3_A5 |
|  |  | -7,031 | 2,25 | -3,125 | 0,002 | 0,01 | A3_A6 |
|  |  | -5,775 | 2,212 | -2,61 | 0,009 | 0,049 | A4_A6 |
| *Gemella* | 123,606 | -3,695 | 1,317 | -2,806 | 0,005 | 0,018 | A2_A6 |
| *Granulicatella* | 273,24 | -3,311 | 0,871 | -3,799 | 0 | 0,001 | A2_A6 |
|  |  | -2,614 | 0,883 | -2,962 | 0,003 | 0,013 | A3_A6 |
| *Haemophilus* | 644,243 | 3,895 | 1,217 | 3,199 | 0,001 | 0,015 | A4_A5 |
| Porphyromonas | 579,685 | -4,086 | 1,18 | -3,464 | 0,001 | 0,002 | A2_A6 |
|  |  | -4,363 | 1,212 | -3,601 | 0 | 0,005 | A5_A6 |
| Pseudoxanthomonas | 27,197 | -23,363 | 3,569 | -6,547 | 0 | 0 | A2_A4 |
|  |  | -21,488 | 3,665 | -5,862 | 0 | 0 | A2_A5 |
|  |  | -23,397 | 3,568 | -6,557 | 0 | 0 | A2_A6 |
|  |  | -33,474 | 3,616 | -9,257 | 0 | 0 | A3_A4 |
|  |  | -31,598 | 3,712 | -8,513 | 0 | 0 | A3_A5 |
|  |  | -33,507 | 3,616 | -9,267 | 0 | 0 | A3_A6 |
| Rothia | 732,239 | -3,995 | 1,024 | -3,903 | 0 | 0,002 | A2_A4 |
|  |  | -4,276 | 1,024 | -4,178 | 0 | 0 | A2_A6 |
| Serratia | 13831,693 | 5,5 | 0,897 | 6,128 | 0 | 0 | A2_A5 |
|  |  | 8,049 | 0,874 | 9,207 | 0 | 0 | A2_A6 |
|  |  | 5,314 | 0,909 | 5,849 | 0 | 0 | A3_A5 |
|  |  | 7,863 | 0,886 | 8,879 | 0 | 0 | A3_A6 |
|  |  | 6,583 | 0,897 | 7,335 | 0 | 0 | A4_A5 |
|  |  | 9,132 | 0,874 | 10,446 | 0 | 0 | A4_A6 |
|  |  | 2,549 | 0,898 | 2,838 | 0,005 | 0,049 | A5_A6 |
| Sphingobacterium | 5,88 | -23,834 | 4,517 | -5,277 | 0 | 0 | A2_A6 |
|  |  | -26,488 | 4,576 | -5,788 | 0 | 0 | A3_A6 |
|  |  | -26,58 | 4,517 | -5,885 | 0 | 0 | A4_A6 |
|  |  | -26,432 | 4,642 | -5,695 | 0 | 0 | A5_A6 |
| Sphingomonas | 930,798 | -2,85 | 0,929 | -3,067 | 0,002 | 0,023 | A2_A5 |
|  |  | -4,278 | 0,904 | -4,73 | 0 | 0 | A2_A6 |
|  |  | -3,282 | 0,916 | -3,583 | 0 | 0,002 | A3_A6 |
|  |  | -3,366 | 0,929 | -3,622 | 0 | 0,004 | A4_A5 |
|  |  | -4,794 | 0,905 | -5,3 | 0 | 0 | A4_A6 |
| Streptococcus | 5556,438 | -1,891 | 0,559 | -3,38 | 0,001 | 0,01 | A2_A4 |
|  |  | -2,473 | 0,559 | -4,421 | 0 | 0 | A2_A6 |
| SEVERE COVID-19 SEVERITY |  |  |  |  |  |  |  |
| Genus | baseMean | log2FoldChange | lfcSE | stat | p-value | Adjusted p-value | Age groups compared |
| *Dolosigranulum* | 551,807 | -24,649 | 5,387 | -4,576 | 0 | 0 | A2_A3 |
|  |  | -17,892 | 5,093 | -3,513 | 0 | 0,01 | A2_A4 |
|  |  | -26,027 | 5,015 | -5,189 | 0 | 0 | A2_A5 |
|  |  | -26,194 | 4,755 | -5,508 | 0 | 0 | A2_A6 |
| *Moraxella* | 31,491 | 24,709 | 5,075 | 4,868 | 0 | 0 | A2_A4 |
|  |  | 19,502 | 4,941 | 3,947 | 0 | 0,003 | A3_A4 |
|  |  | -25,688 | 4,516 | -5,689 | 0 | 0 | A4_A5 |
|  |  | -16,415 | 4,25 | -3,862 | 0 | 0,005 | A4_A6 |
| *Mycoplasma* | 17,595 | 9,358 | 2,561 | 3,655 | 0 | 0,006 | A4_A5 |
| *Shuttleworthia* | 41,453 | -19,204 | 5,39 | -3,563 | 0 | 0,008 | A2_A3 |
|  |  | -16,963 | 5,099 | -3,327 | 0,001 | 0,013 | A2_A4 |
|  |  | -23,964 | 5,016 | -4,778 | 0 | 0 | A2_A5 |
|  |  | -16,521 | 4,768 | -3,465 | 0,001 | 0,011 | A2_A6 |

